# Transmission of SARS-CoV-2 following exposure in school settings: experience from two Helsinki area exposure incidents.

**DOI:** 10.1101/2020.07.20.20156018

**Authors:** Timothee Dub, Elina Erra, Lotta Hagberg, Emmi Sarvikivi, Camilla Virta, Asko Järvinen, Pamela Österlund, Niina Ikonen, Anu Haveri, Merit Melin, Timo Lukkarinen, Hanna Nohynek

## Abstract

**Background:** The role of children in SARS-CoV-2 transmission is unclear. We investigated two COVID-19 school exposure incidents in the Helsinki area.

**Methods:** We conducted two retrospective cohort studies after schools exposures, with a household transmission extension. We defined a case as an exposed person with either a positive RT-PCR, or positive microneutralisation testing (MNT) as confirmation of SARS-CoV-2 nucleoprotein IgG antibodies detection via fluorescent microsphere immunoassay (FMIA). We recruited close school contacts and families of school cases, calculated attack rates (AR) on school level and families, and identified transmission chains.

**Findings:** In incident A, the index was a pupil. Participation rate was 74% (89/121), and no cases were identified. In incident B, the index was a member of school personnel. Participation rate was 81% (51/63). AR was 16% (8/51): 6 pupils and 1 member of school personnel were MNT and FMIA positive; 1 pupil had a positive RT-PCR, but negative serology samples. We visited all school cases’ families (n=8). The AR among close household contacts was 42% (9/20 in 3/8 families) but other plausible sources were always reported. At three months post-exposure, 6/8 school cases were re-sampled and still MNT positive.

**Interpretation:** When the index was a child, no school transmission was identified, while the occurrence of an adult case led to a 16% AR. Further cases were evidenced in 3 families, but other transmission chains were plausible. It is likely that transmission from children to adults is limited.

**Funding:** The Finnish Institute for Health and Welfare funded this study.

**Research in context:** *Evidence before the study:* The first autochthonous case of COVID-19 in Finland was identified on February 29^th^. Transmission of the virus has led to more than 7250 cases and over 300 deaths (As of July 12^th^ 2020). On March 16^th^, assuming that children might have a role in transmission, the Finnish government ordered school closures, to the exclusion of pre-school and grades 1-3. Schools were closed from March 18 and reopened on May 14^th^. At the stage of closure, a very limited number of reports of school related COVID-19 clusters or exposure incidents had been published, and the potential extent of transmission in a school setting was unknown.

*Added value of this study:* We investigated two exposure incidents in two different schools from the Helsinki area to assess transmission among pupils, school personnel and household contacts of identified cases. In school A, contact with a COVID-19 pupil did not lead to further transmission, while in school B, out of 51 recruited contacts, eight (16%) were proved to have had COVID-19 infection, including one member of staff. Among the close household contacts of pupils who were tested positive, COVID-19 attack rate was 31% (5/16). However, in all investigated households, other sources of infections were plausible; hence household transmission following a pediatric COVID-19 case appears to be limited.

*Implications of all of the available evidence:* Incidence of COVID-19 infections in children following school related exposure was limited, as well as secondary transmission within their household. We hope our findings will help prioritize mitigation measures as well as reduce worry among parents of school aged children as most EU countries are preparing for the start of a new school year in autumn.

## Introduction

Since the emergence of COVID-19 in late December 2019 and its development into a pandemic, children appear less likely to develop severe infection or suffer from fatal outcomes.^1,2^ However, it was initially assumed that children could fuel transmission in general population, as is the case with influenza.^3^ Hence many countries, including Finland, ordered temporary school closures as an additional control measure to reduce SARS-CoV-2 transmission.

### COVID-19 in Finland: timeline of events

In Finland, the first imported COVID-19 case was detected in a Chinese tourist on January 29^th.^ and the first autochthonous case on February 29^th^.^4^ On March 12^th^, the Finnish government banned gatherings of more than 500 people and recommended remote working before declaring state of emergency on March 16^th^ : ordering school closures (to the exception of pre-school and grades 1-3: where parents were advised to keep their children at home), advising non-essential businesses to close and banning gatherings of more than 10 people. There was no strict lockdown, but elderly were strongly advised to stay home. On March 27^th^: the Uusimaa region (including Helsinki) was quarantined from the rest of the country (except for essential travel) until April 15^th^. On May 14^th^, schools were re-opened.

### Description of the exposure incidents

#### Exposure incident A

Early March 2020, a 12 year-old pupil from school A was diagnosed with COVID-19. Onset had occurred late February and the pupil had been experiencing minor symptoms while attending school and a team sport training during one day. The training was held outdoors.

Following confirmation of diagnosis, we identified 121 close school and sports contacts: 103 school close contacts (96 pupils from four separate classes and 8 members of school personnel) and 18 close sports contacts: 2 adults and 16 children. One child was identified both as a school and sports contact; for the purpose of the analysis, he was classified as a close sports contact. All close contacts were ordered to remain in home isolation for 14 days starting from the day of exposure and contact healthcare in case of symptom onset.

#### Exposure incident B

In March 2020 a middle aged member of personnel of school B was diagnosed with COVID-19. The person had been experiencing symptoms four to five days earlier and attended work for two days while symptomatic before self-isolating oneself. Contact tracing identified 63 exposed persons: 52 pupils (two classes) and 11 members of personnel. They were all ordered to remain in home isolation for 14 days and contact healthcare in case of symptom onset.

#### Aim of the study

Following these two COVID-19 exposure incidents, we designed a retrospective cohort study of school contacts. We also examined the household contacts of the school-related secondary cases. Our aim was to assess COVID-19 transmission among children, proportion of asymptomatic cases, and study the extent of household transmission, once a child attending the school had been found positive.

## Methods

### Procedures

In exposure A, we collected nasopharyngeal and serum specimens from all participants. In exposure B, as close contacts were invited to participate more than 28 days after exposure, we only collected serum specimens. Short questionnaires were collected from all exposure A and exposure B participants. The household members of participants with a positive test, were invited to participate to the household transmission study and give serum samples. We used a Finnish adaptation of the World Health Organization Household transmission investigation protocol for 2019-novel coronavirus (COVID-19) infection questionnaire.^5^

School B close contacts were also invited for a second serum specimen collection, more than three months after the exposure incident.

### Operational definitions

In both exposure A and B, school contacts included pupils who attended the same classes or shared the same lobby. Teachers of the respective classes and other school personnel in close contact (< two meters, for at least 15 minutes) with the index case were included as close contacts.

In exposure A, sports team contacts comprised all team mates who attended the practice and coaches who were in close contact (< two meters, for at least 15 minutes) with the index case.

A school-related secondary case was defined as a close contact participating in the study with a PCR confirmation of SARS-CoV-2 infection, whether it was collected through participation to the study or standard healthcare or with antibodies detected by both microneutralisation (MNT) and fluorescent microsphere immunoassay (FMIA) tests.

For the purpose of the investigation of household transmission, we defined several categories of school related cases ‘contacts:

- Household contacts
  - A close household contact, i.e. an individual sharing the main residence of the secondary case
  - A regular household contact, i.e. an individual who would regularly host or stay in the same residence of a secondary case (step-sibling, divorced parent and new partner)
- An extended contact, i.e. an individual who would have frequent contact with the secondary case around and after the exposure, for example, grandparents who were involved in caring of the secondary case, according to parents’ reports.

### Microbiological methods

#### Reverse transcriptase Polymerase Chain Reaction

RNA was extracted from specimens with Qiagen Qiacube^R^ instrument using RNeasy Mini Kit^R^. cDNA was synthesized using random hexamer primers and RevertAid H Minus Reverse Transcriptase. The specimen was tested in three separate real-time polymerase reaction tests using QuantiTect^TM^ Multiplex NoRox PCR Kit. Primers and probes were targeted for the envelope (E), the RNAdependent RNA polymerase (RdRp) and the nucleocapsid (N) genes. The sequences of primers and probes are published in Corman et al.^6^

#### Microneutralisation test

SARS-CoV-2-specific neutralising antibody levels were detected from the serum samples using the cytopathic effect (CPE)-based microneutralisation test (MNT) as previously described.^4^ MNT was performed using hCoV-19/Finland/1/2020 (GISAID accession ID EPI_ISL_407079) and hCoV-19/Finland/FIN-25/2020 (EPI_ISL_412971) viruses imported to Finland from China and Italy, respectively. A MNT titer of ≥ 6 was considered as positive.

#### Fluorescent microsphere immunoassay

IgG antibodies to SARS-CoV-2 nucleoprotein (The Native Antigen Company, United Kingdom) were measured with a fluorescent bead-based immunoassay (manuscript in preparation). Antigen was conjugated on MagPlex Microspheres and bound IgG antibodies were identified by a fluorescently labeled conjugated antibody (R-Phycoerythrin-conjugated Goat Anti-Human IgG, Jackson Immuno Research, USA). The plate was read on Luminex® MAGPIX® system. xPONENT software version 4.2 (Luminex®Corporation, Austin, TX) was used to acquire and analyze data. Median fluorescent intensity was converted to U/ml by interpolation from a 5-paramenter logistic standard curve. The specificity and sensitivity of the assay was assessed by plotting a receiver operating characteristic (ROC) curve using the statistical software (GraphPad Prism 8). Antibody concentration equal to or higher than 3·4 U/ml was considered positive, with estimated 100% specificity (Wilson/Brown 95% CI 99·1 to 100%) and 97·9% sensitivity (95% CI 89·1 to 99·9%) of the assay.

### Statistical analysis

We calculated attack rates and described cases using absolute numbers and proportions for categorical variables and median values and interquartile ranges for numeric variables. The initial investigation data was entered using EpiData (version 4.6.0.2), while we used an in-house web-based platform for data entry of the household study questionnaires. Statistical analysis was performed using Stata 15 (StataCorp LP Lakeway, TX, USA), while transmission chains were designed using the R package epicontacts.^7^

### Ethical requirements

The Finnish communicable diseases law and the law on the duties of the Finnish Institute for Health and Welfare allowed the implementation of this research without seeking further institutional ethical review.^8,9^ Informed consent was obtained from all cases and contacts willing to participate in the investigation, before any procedure was performed as part of the investigation. Consent for children under the legal age of consent (15 years) was obtained from a parent or legal guardian. An informed consent was also obtained from the children under the age of 15 years.

## Results

### Exposure incident A

All close contacts (n=121) were invited to come to a healthcare centre in vicinity of school A and/or on the sport training site on day 14 (as quarantine was over) and day 15 after exposure. Participation rate was 74% (89/121) with 74/95 pupils and 15/18 training contacts recruited. Out of 82 nasopharyngeal specimens none were found to be positive. None of the 87 serum samples had detectable SARS-CoV-2-specific neutralizing antibodies nor IgG antibodies to SARS-CoV-2 nucleoprotein. Detailed results and description of participants per exposure groups is presented in Table 1. Since there were no secondary cases among those exposed, no further home visits were made.

**Table 1:**
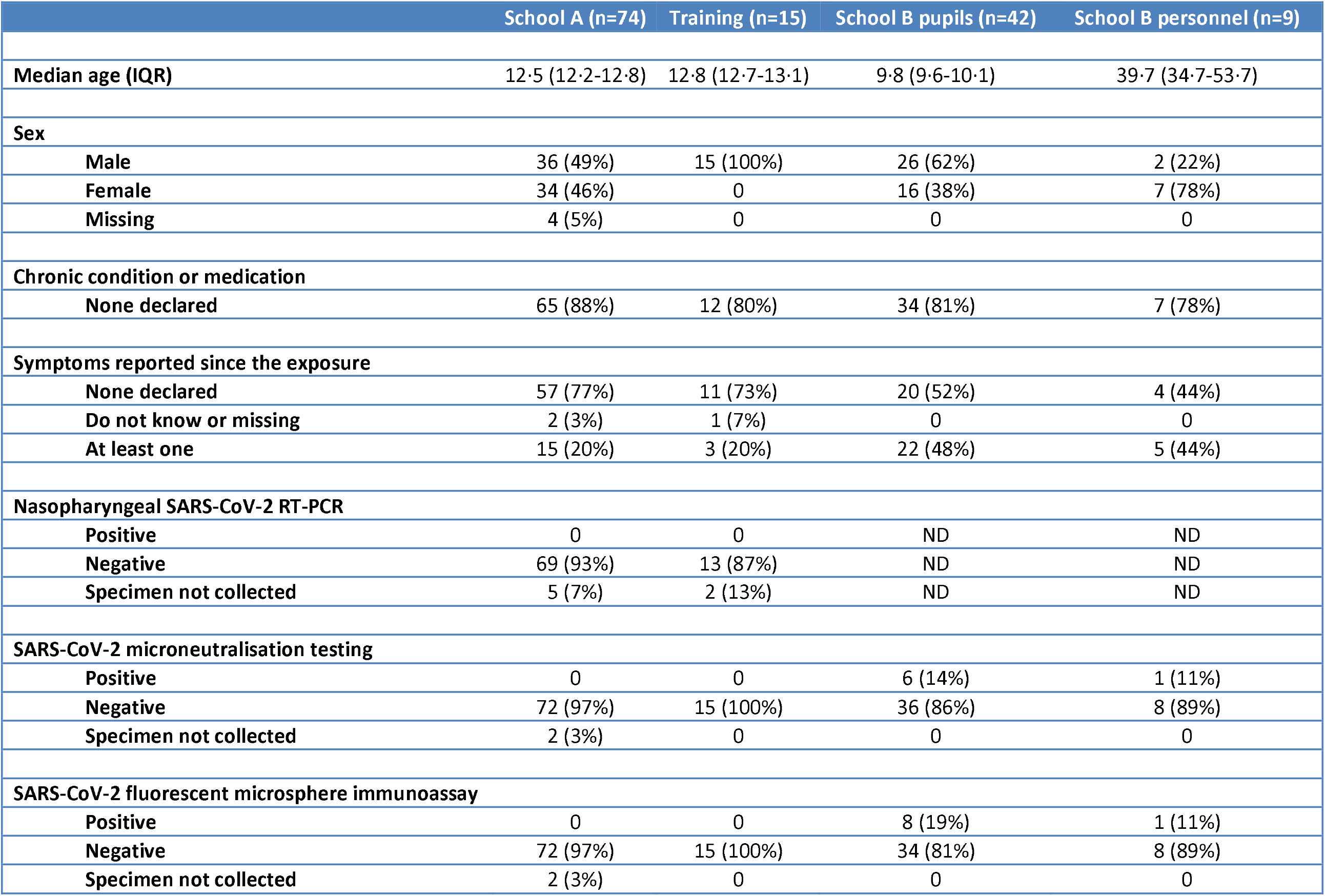
Description of exposure incidents participants.

### Exposure incident B

#### Initial investigations

All pupils (n=52) and school personnel (n=11) that were identified as exposed were invited for collection of serum specimen and questionnaires, more than 28 days after exposure. Participation rate was 81% (42/52) among pupils and 82% (9/11) among school personnel. Detailed results and description of participants per exposure groups is presented in Table 1.

Out of 42 pupils, 6 (14%) were found to have neutralizing antibodies and IgG antibodies to SARS-CoV-2 nucleoprotein, while 2 pupils had low levels (3·6 and 8·7 U/ml) of IgG antibodies to SARS-CoV-2 nucleoprotein measured by FMIA in the absence of neutralizing antibodies, hence were not considered as cases. Additionally, one pupil with a documented PCR confirmation of COVID-19 infection 41 days prior to serum sample collection was tested negative in microneutralisation and FMIA tests. Hence, in total, the secondary attack rate among school B pupils was 17% (7/42). Out of 9 members of school personnel, only one had neutralizing antibodies as well as IgG antibodies to SARS-CoV-2 nucleoprotein (11%). Clinical description of school B related cases is presented in Table 2.

**Table 2:** clinical description of school B secondary cases.

|  | Pre-existing condition | MNT results |  | FMIA test results (U/ml) | PCR results | Symptoms and date of onset |
| --- | --- | --- | --- | --- | --- | --- |
|  |  | Fin-1-20 (titer) | Fin-25-20 (titer) |  |  |  |
| **Pupil 1** | None | 64 | 64 | 26 | Positive on day 7 post exposure | None |
| **Pupil 2** | None | 128 | 64 | 24 | Positive on day 25 post exposure | Day 21 post exposure: Fever, respiratory symptoms |
| **Pupil 3** | None | 64 | 32 | 29 | Not tested | None |
| **Pupil 4** | None | Negative | Negative | Negative | Positive on day 6 post exposure | Day 2 post exposure: Cough |
| **Pupil 5** | None | 64 | >32 | 36 | Negative on day 5 post exposure | Day 4 post exposure: Fever, cough, diarrhea |
| **Pupil 6** | None | 48 | 24 | 68 | Not tested | None |
| **Pupil 7** | None | 128 | 32 | 54 | Not tested | 9 days before exposure: Sore throat<br>7 days post exposure : Headache, diarrhea |
| **School personnel 1** | None | 32 | 24 | 4.5 | Not tested | 24 days post exposure: Runny nose, fever |

MNT: SARS-CoV-2-specific neutralising antibody test
Fin-1-20 (titer): hCoV-19/Finland/1/2020
Fin-25-20: hCoV-19/Finland/FIN-25/2020
FMIA: IgG antibodies to SARS-CoV-2 nucleoprotein detection via fluorescent microsphere immunoassay

Age of pupils considered positive (median: 10·1 [IQR: 9·7-10·1]) was not significantly different from pupils who were considered as negative (median: 9·8 [IQR: 9·6 – 10·0]).

At three months post exposure, 19/52 (37%) pupils and 5/11 (45%) members of school personnel participated to a second collection of serum specimen. No new cases were identified and all previously identified cases who were sampled (5 pupils and 1 member of personnel) were still MNT positive.

#### Household transmission study

We contacted all secondary COVID-19 cases’ families (n=8): seven pupils (6 MNT positive and FMIA positive pupils and 1 pupil with a documented positive PCR but tested MNT and FMIA negative) and one MNT positive and FMIA positive member of personnel.

All cases’ families agreed to participate and samples were collected from 32/33 contacts: 19/20 close household contacts, 9/9 regular household contacts and 4/4 extended household contacts.

In total, secondary household transmission was detected in eight close household contacts out of 19 (42%) with available samples: eight close contacts had both FMIA and MNT positive results (Figure 1), while one close contact was only FMIA positive with low levels (5·4 U/ml) of IgG antibodies to SARS-CoV-2 nucleoprotein, hence not considered as a secondary case. No further transmission was detected in regular household contacts and extended contacts (Table 3). All MNT positive contacts originated from three households.

**Table 3:** Number of contacts and secondary attack rate among contacts of school B related cases.

| Identifier | Close household contacts |  |  | All (close and regular) household contacts |  |  | All (household and extended) contacts |  |  |
| --- | --- | --- | --- | --- | --- | --- | --- | --- | --- |
|  | Sampled | Positive contacts | Secondary attack rate | Sampled | Positive contacts | Secondary attack rate | Sampled | Positive contacts | Secondary attack rate |
| **Pupils** |  |  |  |  |  |  |  |  |  |
| **Pupil 1** | 1 | 0 | 0% | 2 | 0 | 0% | 2 | 0 | 0% |
| **Pupil 2** | 3 | 3 | 100% | 3 | 3 | 100% | 3 | 3 | 100% |
| **Pupil 3** | 2 | 0 | 0% | 4 | 0 | 0% | 4 | 0 | 0% |
| **Pupil 4** | 3 | 0 | 0% | 3 | 0 | 0% | 3 | 0 | 0% |
| **Pupil 5\*** | 2 | 0 | 0% | 5 | 0 | 0% | 7 | 0 | 0% |
| **Pupil 6** | 3 | 0 | 0% | 3 | 0 | 0% | 3 | 0 | 0% |
| **Pupil 7** | 2 | 2 | 100% | 5 | 2 | 40% | 7 | 2 | 29% |
| **Total** | 16 | 5 | 31% | 25 | 5 | 20% | 29 | 5 | 17% |
| **School personnel 1** | 3 | 3 | 100% | 3 | 3 | 100% | 3 | 3 | 100% |
| **Total** | 19 | 8 | 42% | 28 | 8 | 29% | 32 | 8 | 25% |

\*One close contact did not wish to participate to serum collection

**Figure 1:**
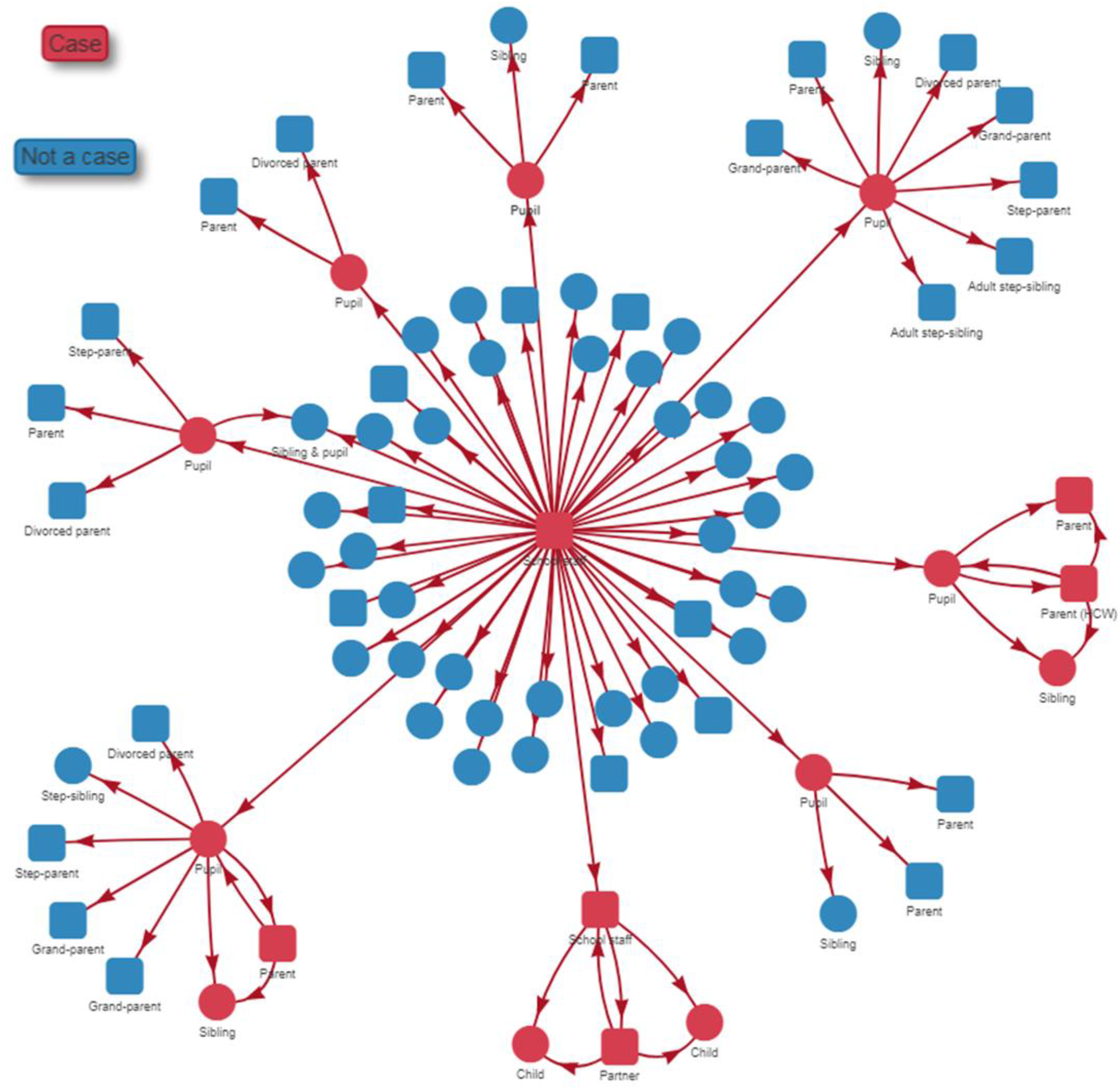
School B exposure incident transmission chains. Case defined as participant with either PCR confirmation of SARS-CoV-2 infection or with antibodies detected by both microneutralisation (MNT) and fluorescent microsphere immunoassay (FMIA) tests. Participants aged less than 18 years old are displayed using rounds, while participants aged 18 or more are represented with squares.

In household 2, the pupil had experienced onset of respiratory symptoms and fever, respectively at 21 and 22 days after exposure. At the same time, one of his parents who is a healthcare worker, reported respiratory symptoms for a two-week period and loss of taste for a couple of days, and was PCR-confirmed. As symptom onset occurred more than 14 days after exposure at school, and due to the parent’s profession, it is very likely that COVID-19 transmission in this family was not school related. The other MNT positive parent also reported respiratory symptoms, fever, headache, and muscle pain during the same time period, while the younger sibling only had had fever for one day.

In household 7, the pupil had experienced different symptoms at two different timepoints: a sore throat starting at nine days before exposure; headache and diarrhea starting at seven days post exposure. His two close contacts both were IgG positive and had SARS-CoV-2 specific neutralizing antibodies: an asymptomatic younger sibling and a parent who reported to have had sore throat, headache and fatigue over a period of 19 days starting at five days before the school exposure incident.

In the household of school personnel 1, all contacts were IgG positive and were found to have SARS-CoV-2-specific neutralising antibodies. Questionnaire data showed that the school personnel 1’s partner started to have fever 21 days after the school exposure incident, a few days before school personnel 1 got sick (24 days post exposure)Their children only reported fever on the day after school personnel 1 got ill. As the exposed person fell ill more than 2 weeks after the school exposure and as one of the family members was symptomatic earlier, it is possible that this family cluster was not school related, similarly as in household 2.

## Discussion

We studied two school exposure cases and found that exposure to a 12-year-old case with mild symptoms led to no further transmission of SARS-CoV-2, whereas exposure to an adult school staff member with moderate symptoms led to secondary transmission.

Earlier reports on school exposure with a child as the index case show variable results. In France, Danis et al. studied 54 school contacts who, based on nasopharyngeal PCR-testing, showed no further transmission.^10^ In Ireland, surveillance over a 12-day-period showed that out of 1,160 PCR confirmed cases, only six cases had history of school attendance and no further confirmed COVID-19 cases were detected among their 1025 contacts: 924 children and 101 adults.^11^ Still, in New South Wales (Australia), following the detection of 18 COVID-19 cases, only two pupils out of 863 close contacts were identified as secondary cases either via nasopharyngeal PCR testing or detection of antibodies in a serum sample.^12^

In the French region of Oise, the investigation of a high-school cluster, with majority of pupils aged 15-17 years, used three different methods: an ELISA, a flow-cytometry and an immunoprecipitation assay to show that the attack rate was 38% (92/240). This attack rate was twofold higher than the 17% we found in school B. However, the fact that pupils were older than in our exposure incident might explain this difference. In this same investigation, secondary household transmission rate was 19% (23/121), which was in line with our findings^13^. On the other hand, in the same region, a study conducted among primary school aged children (6 – 11 years old) showed a 9% attack rate (45/510) among pupils, with almost half of the cases being asymptomatic.^14^ It appears plausible that infectiousness and/or susceptibility would differ with age.

Bi et al. in their study on transmission of COVID-19, using nasopharyngeal RT-PCR in 391 cases and 1286 of their close contacts, did not see a difference in the secondary attack rate depending on the age of the index case.^15^ Neither did a Spanish nationwide seroprevalence study find major difference between children and young adults.^16^ Xu et al. reported that out of 9,120 Chinese confirmed cases in January-February 2019, transmission from primary cases to others was lower in children aged 0-17 years than with older index cases.^17^

We only found one previous report where the index case in a school setting would have been an adult leading to a cluster of 16 adult staff members, but no cases among preschool pupils (median age: 4 years old) diagnosed by PCR.^18^ Here, we found that an adult index case could lead to further transmission among children with the identification of eight secondary cases among pupils and one among school personnel. Age might have a role in susceptibility to SARS-CoV-2 infection in children. However, as symptom onset was quite late in several cases and as other plausible transmission chains were often identified, it is likely that our findings are overestimating the attack rate after school exposure. Additionally, when we collected further serum samples two months after the initial collection, after re-opening the schools, none of the 18 previously negative participants that were sampled again had seroconverted.

In Finland, access to PCR diagnosis testing was restricted until middle of April, hence it is plausible that in general population, pediatric cases and possible school-related transmission might have been missed. However, as testing capacity rapidly increased, and access to testing was good, we did not see an increase in cases among children after reopening the schools for a two-week period either.^19^

None of the nasopharyngeal samples were positive during the initial investigation at school A. The method is both operator and time dependent, and some cases might have been missed, but as we did not find any MNT positive or FMIA positive among participants, it is unlikely that we would have missed all the cases.

When serology is used to assess previous COVID-19 infection, the sensitivity and the specificity of the used antibody test is critical. With tests that measure binding antibodies, sometimes cross-reactive antibodies (e.g., induced by prior infections with seasonal coronaviruses) may cause false positive test results. Neutralizing antibodies are considered a highly specific indication of a prior COVID-19 infection. Therefore, in this study a case was defined primarily by a positive MNT result. All cases with positive MNT results also had elevated IgG concentrations to SARS-CoV-2 nucleoprotein. We used a stringent cut-off value for seropositivity in the FMIA antibody test to minimize the chance of false positive test results. Serum samples that tested positive for IgG but negative for MNT had low antibody levels close to the threshold value, which may reflect the presence of cross-reactive antibodies. It has also been suggested that not all subjects who are infected with COVID-19 develop neutralizing antibodies, especially patients with mild symptoms. A study of COVID-19 cases with mild symptoms, detected neutralizing antibodies in 79%, 92% and 98% of samples collected on days 13-20, 21-27 and 28-41 after symptom onset, respectively.^20^ Another study found that 10/175 patients with mild symptoms did not produce neutralizing antibodies even during a longer follow-up.^21^ Some studies have found neutralizing antibodies in all cases, even in those with mild symptoms.^22,23^ This suggests that most if not all subjects with COVID-19 develop neutralizing antibodies eventually, and thus using a serological test based on measuring neutralizing antibodies can be considered as a very sensitive and specific method for assessing prior COVID-19 infection, when serology is assessed at late convalescent phase. In this study we also found that neutralizing antibodies persisted at least for two months after initial sample collection.

Finally, a limitation of this work is the rather small sample size: 89 participants in exposure incident A and 51 in incident B, however, the 76% participation rate obtained in the initial investigation, and 100% in the household transmission study does improve the representativeness of our findings. We were only made aware of one additional COVID-19 case among close contacts who did not participate to the study.

## Conclusion

We found that exposure in school due to a child index case with mild symptoms led to no further cases among the close contacts, whereas exposure to an adult index case with moderate symptoms in a school setting led to secondary transmission. The outcome is most likely a result of several factors including the age of the index case, the degree of symptoms, and the type of contact. Further studies on COVID-19 transmission in a school setting are required in order to better advice on mitigation measures such as quarantine and school closure. Starting from August 2020, the Finnish Institute for Health and Welfare will be coordinating multisite transmission studies in the five university hospital cities to further understand the role of children in SARS-CoV-2 transmission chains and the role of immunity and other factors in this process.

## Data Availability

Data cannot be transferred or shared

## Authors’ contributions

TD, ES, NI, MM, and HN conceived and designed the study. EE, AJ and TL conducted contact tracing. TD, LH, ES, and HN recruited participants to the study. NI developed, supervised and interpreted the virological analysis, PÖ cultivated the viral strains for immunological analyses. CV, AH, and MM developed, performed, supervised and interpreted the immunological analyses. TD performed the statistical analyses. TD and EE wrote the first version of the report. All authors critically reviewed and approved the final version of the report

## Acknowledgements

We are grateful to the skillful technical assistance of HUSLAB and THL staff for their technical assistance in the lab, as well as both school administrative personnel for their help in organizing recruitment of participants. We also wish to thank Isabel Bergeri and Rebecca Grant from the World Health Organisation Emergencies Programme for their input on our protocols.

